# Long-COVID improves in 50% of patients after a year in a Midwestern cohort

**DOI:** 10.1101/2024.04.30.24306497

**Authors:** Grant Stalker, Rosarie Tudas, Alpana Garg, Lauren Graham, Andrew L. Thurman, R. Todd Wiblin, Nabeel Hamzeh, Robert J. Blount, Raul Villacreses, Joseph Zabner, Alejandro Comellas, Josalyn L. Cho, Alejandro Pezzulo

## Abstract

**Background:** Many of those infected with COVID-19 experience long-term disability due to persistent symptoms known as Long-COVID, which include ongoing respiratory issues, loss of taste and smell, and impaired daily functioning.

**Research Question:** This study aims to better understand the chronology of long-COVID symptoms.

**Study Design and Methods:** We prospectively enrolled 403 adults from the University of Iowa long-COVID clinic (June 2020 to February 2022). Participants provided symptom data during acute illness, symptom progression, and other clinical characteristics. Patients in this registry received a survey containing questions including current symptoms and status since long-COVID diagnosis (sliding status scale, PHQ2, GAD2, MMRC). Those >12 months since acute-COVID diagnosis had chart review done to track their symptomology.

**Results:** Of 403 participants contacted, 129 (32%) responded. The mean age (in years) was 50.17 +/−14.28, with 31.8% male and 68.2% female. Severity of acute covid treatment was stratified by treatment in the outpatient (70.5%), inpatient (16.3%), or ICU (13.2%) settings. 51.2% reported subjective improvement (sliding scale scores of 67-100) since long-COVID onset. Ages 18-29 reported significantly higher subjective status scores. Subjective status scores were unaffected by severity. 102 respondents were >12 months from their initial COVID-19 diagnosis and were tracked for longitudinal symptom persistence. All symptoms tracked had variance (mean fraction 0.58, range 0.34-0.75) in the reported symptoms at the time of long-COVID presentation when compared with patient survey report. 48 reported persistent dyspnea, 23 (48%) had resolved it at time of survey. For fatigue, 44 had persistence, 12 (27%) resolved.

**Interpretation:** Overall, 51.2% respondents improved since their long-COVID began. Pulmonary symptoms were more persistent than neuromuscular symptoms (anosmia, dysgeusia, myalgias). Gender, time since acute COVID infection, and its severity didn’t affect subjective status or symptoms. This study highlights recall bias that may be prevalent in other long-COVID research reliant on participant memory.

## Introduction

Beginning in 2020, some patients recovering from COVID-19 noticed persistent symptoms well past the acute infection phase. Termed long-COVID or post-acute sequelae SARS-CoV-2 infection (PASC), the condition is characterized by a variety of symptoms including fatigue, dyspnea, myalgias, headache, and difficulty concentrating among others (1-3). Long-COVID has similarities when compared to other conditions (i.e. chronic fatigue syndrome (4)), however the risk factors, clinical presentation, and time course are incompletely known. An obstacle to accurately characterizing long-COVID has been the lack of cohorts that include both mild and severe COVID-19 and includes patients from rural settings. In this study, we combined a survey with data obtained from participant charts to determine risk factors, prevalence, and clinical time course of long-COVID symptoms. We hypothesized that long-COVID duration would correlate with acute disease severity and with participant demographics. As shown below in results, we found that younger age at diagnosis correlated with higher likelihood of improvement and that at approximately one year (1.45 ± 0.47 years) after initial clinic visit, nearly half of patients experienced significant overall symptom improvement or resolution.

## Methods

### Cohort

We prospectively enrolled 403 adults (age 18 or older) with long-COVID who were initially evaluated in the University of Iowa long-COVID clinic between June 2020 and February 2022 (5). Participants were defined as having long-COVID if any symptom persisted more than 30 days after a positive rapid antigen or reverse transcriptase–polymerase chain reaction test from a nasopharyngeal or oropharyngeal swab or a positive SARS-CoV-2 nucleocapsid serum antibody test.

Clinical data was extracted from the electronic medical record (EMR) by two researchers who reviewed outpatient, emergency department, and hospitalization records from the University of Iowa and regional hospitals to collect data. The period of acute infection with SARS-CoV-2 was defined as 21 days following a positive COVID-19 diagnosis and symptoms that persisted past this period that were unattributed to other causes were considered long-COVID symptoms (6). This study is approved by the University of Iowa Institutional Review Board, Study #202204252.

### Data Collection

Participants in the overall cohort underwent extensive standardized collection of clinical, imaging, and laboratory data, including a list of symptoms assessed in every patient. For the purposes of this study, we analyzed demographic data, symptoms and disease severity during acute illness, and symptoms reported during initial clinic visit.

### Symptom Survey

To better understand the time course of long-COVID symptoms, all participants in the cohort with an available email address were sent a link to a REDCap survey. Patients who did not respond within 2 weeks or who did not have an email received a phone call followed by an email with the survey link if interested in participating. The survey included previously validated [Patient Health Questionnaire 2(PHQ-2), Generalized Anxiety Disorder −2 item (GAD-2), Modified Medical Research Council Dyspnea Scale (MMRC)] tools to assess depressed mood, anxiety, and dyspnea (dyspnea only if symptom present) (7-10). At the time of data collection, no peer-reviewed questionnaires were available for assessment of clinical status in people with long-COVID. Therefore, we also developed a subjective status score utilizing a continuous sliding numerical scale (0 being worse, 50 being no change, 100 being fully recovered at the time of survey response compared to first clinic visit). Additionally, the survey contained questions about vaccine status, reinfection status, and current symptoms, including the same symptoms documented at the initial clinic visit. Patients who responded to the survey over a year after their diagnosis of acute COVID-19 had further chart review done to track their symptomology from the time of their acute COVID-19 infection to their survey response using office visits, emergency room visits, and hospital visits to track symptoms. Persistent symptoms were defined as symptoms that occurred during the acute COVID-19 infection and continued to occur at their first follow up visit for long-COVID symptoms (i.e. patient had dyspnea during their acute COVID-19 infection and continued to have dyspnea at their visits after the 21 day window).

### Comparisons

To assess whether the persistence of long-COVID is associated with patient characteristics or the COVID-19 infection characteristics, participants were grouped according to demographics (sex and age ranges), acute illness severity (mild = outpatient, moderate = inpatients, severe = intensive care unit admission), and time since acute COVID-19 diagnosis to perform data comparisons. Grouping by subjective status was pre-specified as “worse”, “no change” or “improved” according to continuous score of 0-32, 33-66, and 67-100 respectively. The time course of specific symptoms was assessed only in patients that had the persistent symptom at first clinic visit. Patients were compared using the subjective status score, GAD-2, PHQ-2, and MMRC.

### Data Analysis

Continuous variables are reported using means with standard deviations. Categorical variables are reported as counts and percentages. Categorical variables were compared using chi-squared tests and continuous variables were compared using t-tests. Differences were considered statistically significant when P < .05 based on a two-tailed test. To trace symptom persistence over time, survival analysis was performed. Failure times were interval censored. The right bound of the censoring interval was the first clinic visit in which a patient reported the symptom had resolved, and the left bound was the most recent prior clinic visit in which the symptom was present. Kaplan-Meier curves were estimated for each symptom. Statistical analyses were performed using GraphPad Prism (version 9.4) or R (version 3.5.1, survival package version 2.42, http://www.r-project.org).

## Results

We contacted 403 participants and received completed surveys from 129 (32%) (Table 1). The average age of respondents was 50.17 ± 14.23 years. 31.8% of the respondent cohort was male and 68.2% was female. The severity of acute covid treatment was stratified by treatment in the outpatient (70.5%), inpatient (16.3%), or ICU (13.2%) settings. 12.4% (16 respondents) had at least one dose of a COVID-19 vaccine prior to their infection. We compared the respondent cohort to the overall cohort and found similar demographics for each group of participants including gender, age, severity of initial COVID infection, and race (Table 1).

**Table 1:** Survey Respondent Demographics for the overall University of Iowa long-Covid Cohort and survey respondent cohort.

| Overall long-COVID Cohort |  | Survey respondent Cohort |  |
| --- | --- | --- | --- |
| **Gender** |  | **Gender** |  |
| *Male* | 154 (33.33%) | *Male* | 41 (31.78%) |
| *Female* | 308 (66.67%) | *Female* | 88 (68.22%) |
| **Age** |  | **Age (years)** |  |
| *18-29* | 65 (14.10%) | *18-29* | 13 (10.08%) |
| *30-39* | 61 (13.23%) | *30-39* | 16 (12.40%) |
| *40-49* | 133 (28.85%) | *40-49* | 35 (27.13%) |
| *50-59* | 104 (22.56%) | *50-59* | 34 (26.36%) |
| *60-69* | 63 (13.67%) | *60-69* | 20 (15.50%) |
| *70+* | 35 (7.59%) | *70+* | 11 (8.53%) |
| **Severity** |  | **Severity** |  |
| *Outpatient* | 338 (73.16%) | *Outpatient* | 91 (70.54%) |
| *Inpatient* | 78 (16.88%) | *Inpatient* | 21 (16.28%) |
| *ICU* | 46 (9.96%) | *ICU* | 17 (13.18%) |
| **Race/Ethnicity** |  | **Race/Ethnicity** |  |
| *White* | 400 (86.58%) | *White* | 120 (93.02%) |
| *Black* | 14 (3.03%) | *Black* | 3 (2.33%) |
| *Hispanic* | 37 (8.01%) | *Hispanic or Latino* | 6 (4.65%) |
| *Other* | 11 (2.38%) | *Other* | 0 (0.00%) |

51.2% of participants reported subjective improvement (sliding scale scores of 67-100), 40.3% reported little or no subjective change (subjective status scale scores of 33-66), and 8.5% reported feeling subjectively worse (sliding scale scores of 0-32) than when they first experienced long-COVID. The average score of the PHQ2 scale for the entire cohort was 3.58 ± 1.85 (a score ≥ 3 raises suspicion for major depressive disorder). Participants in the 18-29 age group reported significantly higher subjective status scores compared to all other ages (p = 0.0101), with 12 out of 13 participants reporting subjective improvement from the time they initially experienced long-COVID to the time of survey completion (Figure 1). PHQ2 Scores did not reach clinical significance when comparing the same groups (0.0739) (Figure 1). To rule out differences in improvement by age group due to varying response times, analysis of time to survey response from initial COVID infection was done. There was no correlation between time to response and subjective status score (Supplemental Figure 1A). Males and females reported similar subjective improvement (Supplemental Figure 1B), and no differences were observed in subjective status scores across acute COVID-19 severity groups (Supplemental Figure 1C). There were no age-related differences in GAD2 or MMRC scores (Supplemental Figure 2).

**Figure 1:**
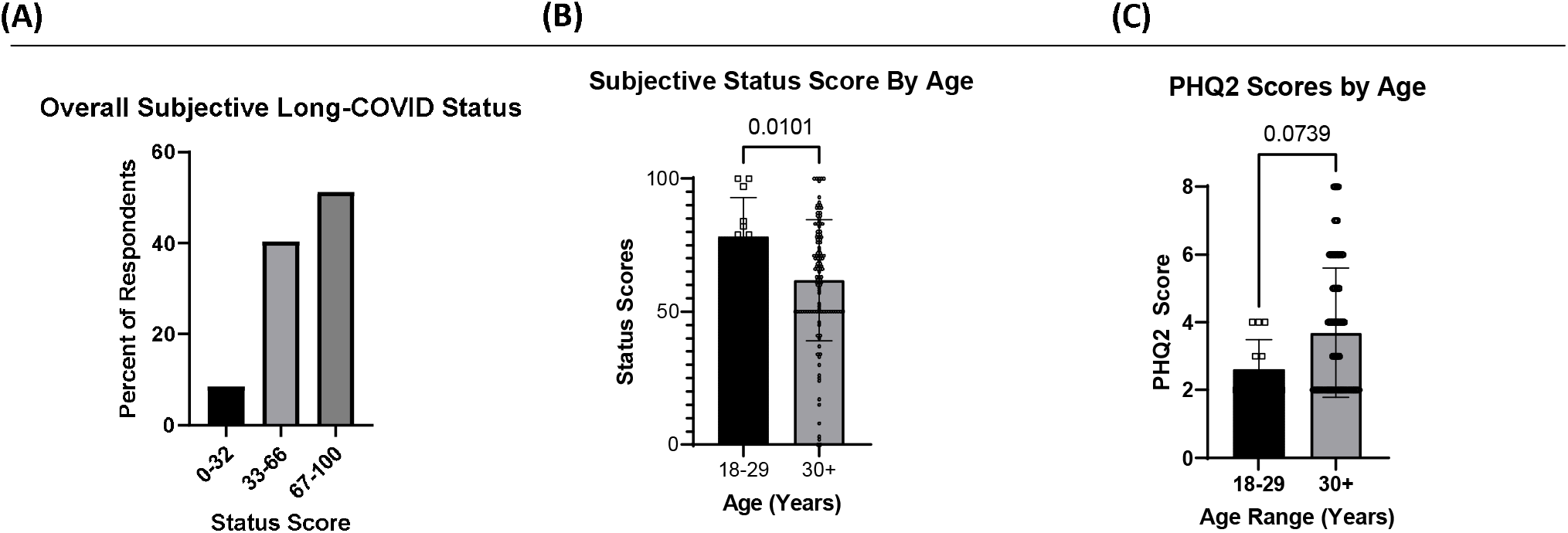
Comparisons of subjective status scores as indicated on survey responses. Mann-Whitney testing used for p-value calculation. **(A)** Overall subjective status scores for all participants who completed the survey. **(B)** Subjective status scores stratified by age. **(C)** PHQ2 Scores at time of survey.

Next, we examined the time course of symptom resolution vs. persistence from the first clinic visit. However, participant recall of specific symptoms is highly affected by recall bias (11-13); therefore, we quantified the concordance between initial clinic visit symptom recall vs. reported in the corresponding clinical note. Symptom discordance was assessed by comparing patient recall of initial symptomatology (acquired via patient survey) to reported symptoms as described in their initial long-COVID clinic note in their EMR. All symptoms tracked had significant discordance (mean fraction 0.58, range 0.34-0.75) in the reported symptoms at the time of long-COVID presentation with patient reported (Supplemental Figure 3). This data suggests that participant recall bias of long-COVID symptoms is high and likely to affect estimates of symptom resolution time course. Since the list of symptoms obtained in the survey matches the standardized list obtained in first clinic visit, we used data from chart review rather than patient recall to estimate resolution or persistence of specific symptoms.

102 respondents were at least 12 months from their initial COVID-19 diagnosis and were tracked for longitudinal symptom persistence (Figure 2). 48 respondents reported persistent dyspnea and 23 (48%) reported recovery of dyspnea during at least one clinical visit or on their survey response. 44 respondents reported persistent fatigue and 12 (27%) reported no fatigue during at least one clinical visit or on their survey response. 21 respondents reported persistent anosmia and 13 (62%) reported no anosmia during at least one clinical visit or on their survey response. 19 respondents reported persistent dysgeusia and 11 (58%) reported no dysgeusia during at least one clinical visit or on their survey response. 22 respondents reported persistent myalgias and 12 (55%) reported no myalgias during at least one clinical visit or on their survey response.

**Figure 2:**
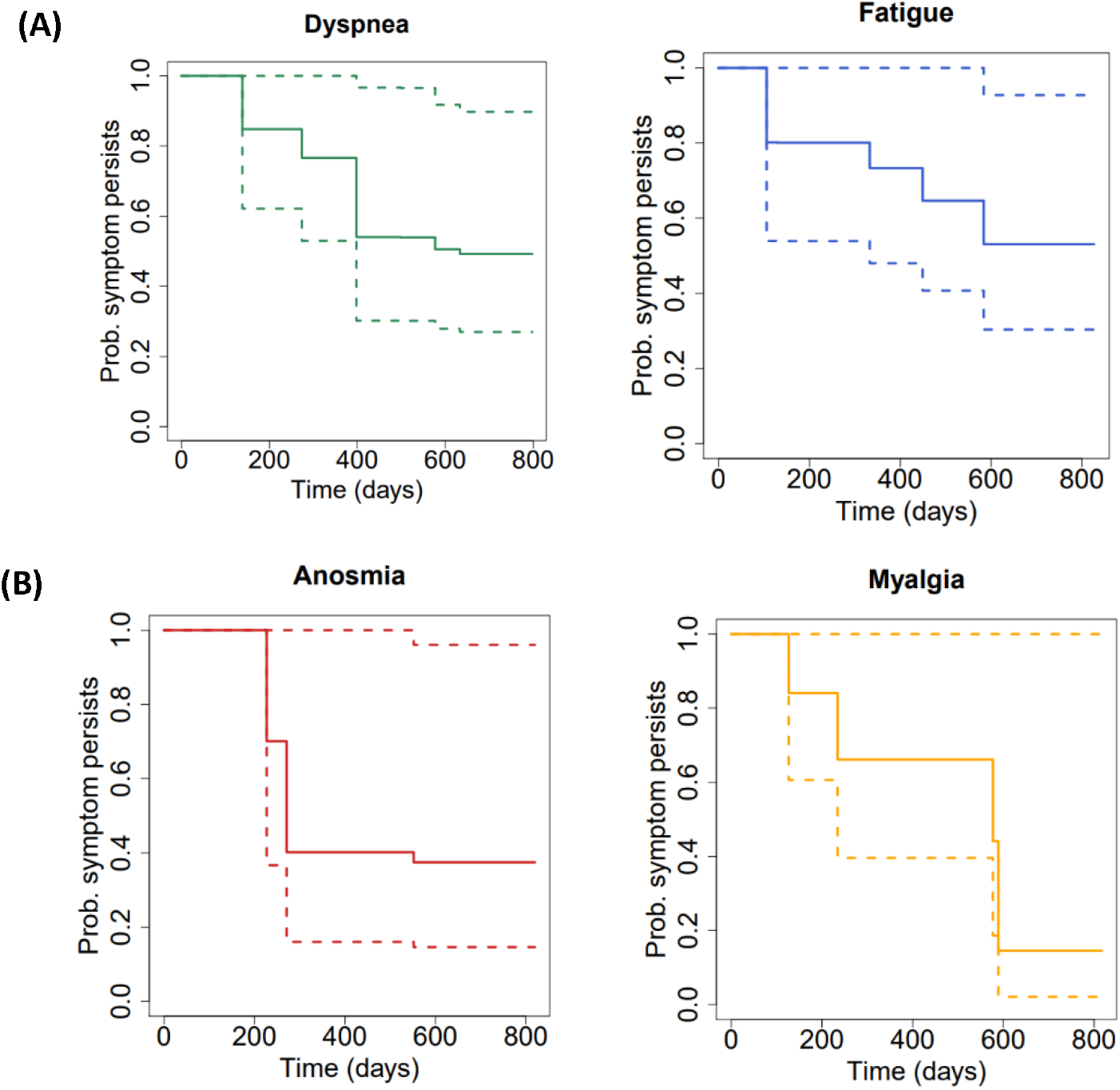
Longitudinal symptom persistence displayed as time to loss of symptoms in days since COVID-19 diagnosis. Dotted lines represent the 95% confidence interval for symptom persistence. **(A)** Likelihood of neuromuscular symptom persistence over time. **(B)** Likelihood of respiratory symptom persistence over time.

## Discussion

In this study we collected data from participants with long-COVID using both survey data and retrospective chart review. Participants were assessed for current symptoms and overall wellbeing, and persistent symptoms were extracted from electronic medical records. Overall, 51.2% of respondents reported an improvement in their overall wellbeing since the onset of long-COVID symptomology. Further, dyspnea was more persistent than anosmia, dysgeusia, and myalgias. Our data suggests that the risk of persistent long-COVID symptomatology is lower in younger people and is consistent with previous data (14). No differences were found in overall wellbeing or specific symptomology in males compared to females or with respect to time since acute-COVID-19 infection or severity of acute-COVID-19.

Inflammation plays a critical role in many disease pathologies with age being a significant factor in increased inflammatory states and markers (15, 16). COVID-19 and long-COVID have been previously shown to cause proinflammatory states and increases in inflammatory cytokines which may play a role in long-COVID symptomology (17, 18). The ability of younger individuals to more quickly resolve inflammation paired with a lower baseline level of chronic inflammation may explain the improvement in symptomology in a younger cohort (19).

This study provides insight into the longevity of long-COVID symptoms and overall course of patient status. Our study design has important strengths. First, participants were evaluated in person during a clinical visit, using standardized data collection tools to facilitate accurate comparisons. Second, the use of survey data and chart review in this study allowed for minimization of the impact of patient recall bias influencing symptom persistence. Recall bias has been well established in medical research impacting a wide range of data collection including symptom recall, dietary recall, and medication use recall (20-22). Interestingly, the similarity between PHQ2 scores during survey collection with the subjective status scores suggest that participant recall of overall wellbeing is less affected by recall bias than recollection of specific symptoms. Third, this study covers a broad range of acute-COVID severities while mirroring the demographics of the state of Iowa, which is a rural population underrepresented in most large cohorts. Finally, the study provides granular data tracking specific symptoms in individuals with long-COVID, which has been poorly characterized to date.

Limitations of this study include the lack of a control group of participants with acute COVID-19 but no long-COVID or participants that never had acute COVID-19. While the unvalidated scale impact was mitigated through use of additional validated measures, further work can look to better characterize long-COVID quality of life with validated questionnaires. Moreover, we did not analyze the correlation between vaccination status/time and symptoms; most first visits occurred prior to approval of the SARS-CoV-2 vaccine. Finally, exact dates for symptom resolution were unobtainable due to study limitations as patients only reported symptoms when seen at follow up visits or when replying to the survey. One important consideration gleaned from this study is the necessity of robust symptom tracking in novel diseases.

## Conclusion

In a cohort of participants with long-COVID, 50% report improvement in wellbeing after a year. This has important implications in caring for patients with long-COVID regarding setting expectations and considering clinical treatment. Future studies will look to standardized follow up and symptom definitions to allow more robust analysis of long-COVID symptomology.

## Data Availability

All data produced in the present study are available upon reasonable request to the authors.

## Acknowledgement

Grant Stalker had full access to all of the data in the study and takes responsibility for the integrity of the data and the accuracy of the data analysis, including and especially any adverse effects. Rosarie Tudas, Alpana Garg, Lauren Graham, Andrew L. Thurman, R. Todd Wiblin, Nabeel Hamzeh, Robert J. Blount, Raul Villacreses, Joseph Zabner, Alejandro Comellas, Josalyn L. Cho, Alejandro Pezzulo contributed substantially to the study design, data analysis and interpretation, and the writing of the manuscript.

## Abbreviation List

GAD-2: Generalized Anxiety Disorder 2-item
PHQ-2: Patient Health Questionnaire-2
MMRC Dyspnea Scale: Modified Medical Research Council
PASC: Post-acute sequelae SARS-CoV-2 infection
EMR: Electronic Medical Record

---

**Supplemental figure 1:**
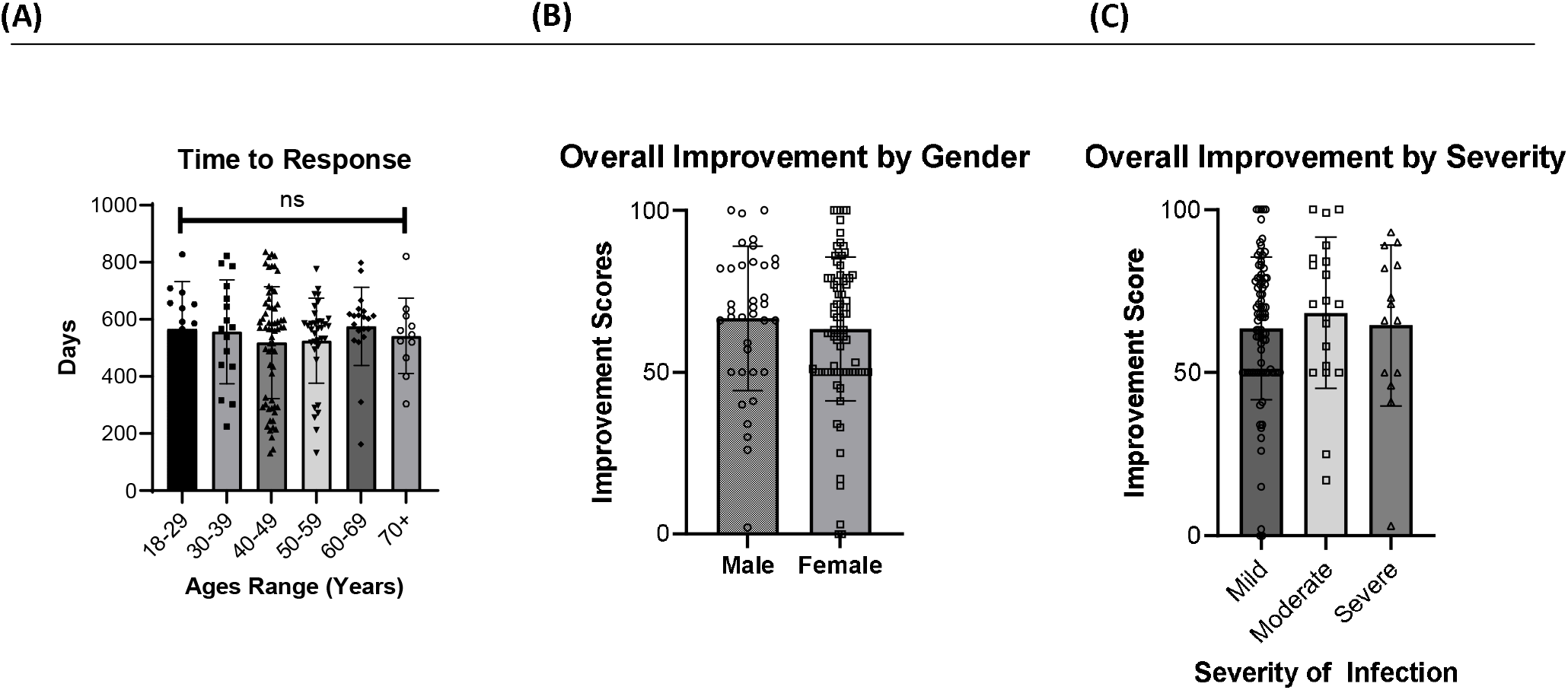
(A) Time to survey response since diagnosis. Overall improvement scores by (B) gender and (C) severity of acute COVID19 infection. ns, non-significant.

**Supplemental figure 2:**
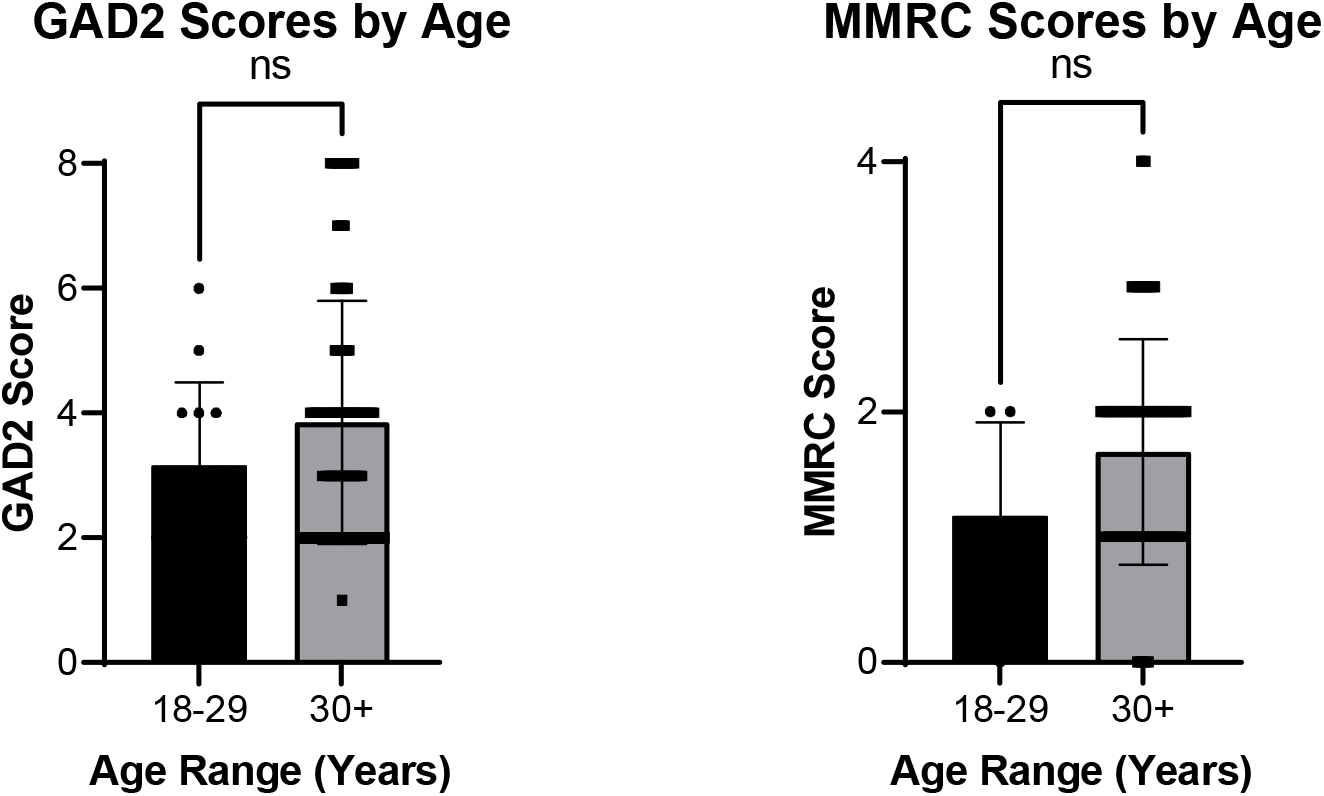
Validated survey scores: **(A)** General Anxiety Disorder 2 item questionnaire at time of survey **(B)** MMRC scores at the time of survey for patients reporting dyspnea

**Supplementary Figure 3:**
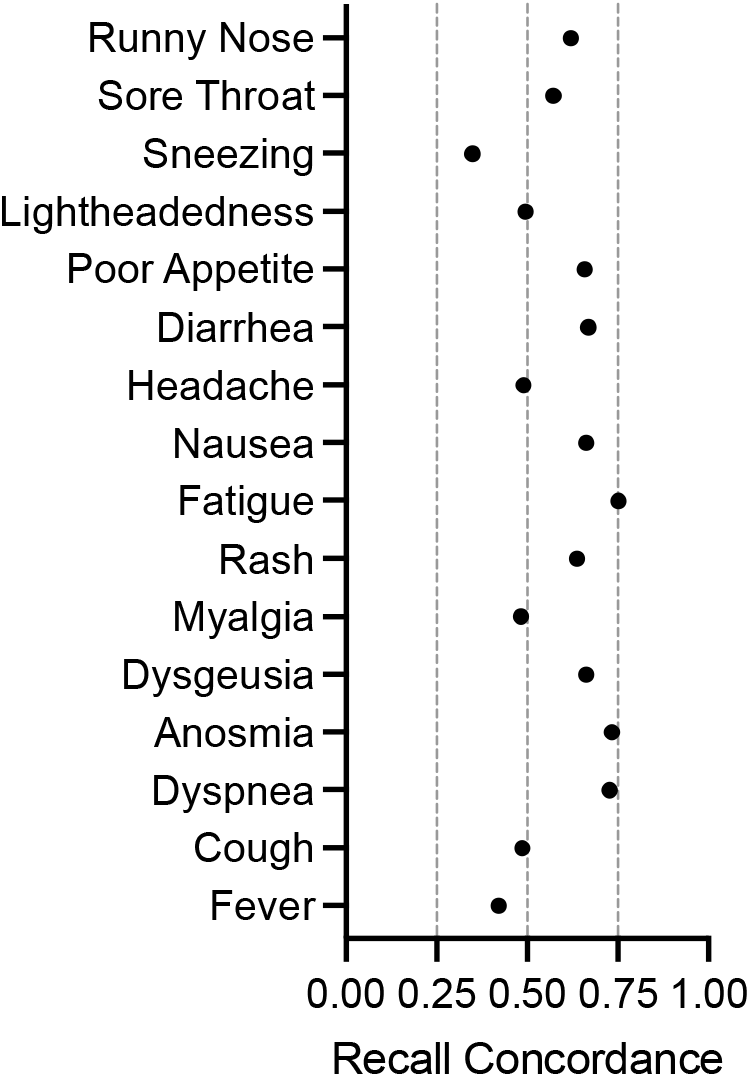
Symptom recall concordance (Patient-reported symptoms, retrospective/recall vs. reported/in chart) for first clinic visit. Data compares symptoms reported at patients’ first long-COVID clinic visit to patients’ recollection of their initial symptoms as indicated on their survey responses.

**Supplemental Figure 4:**
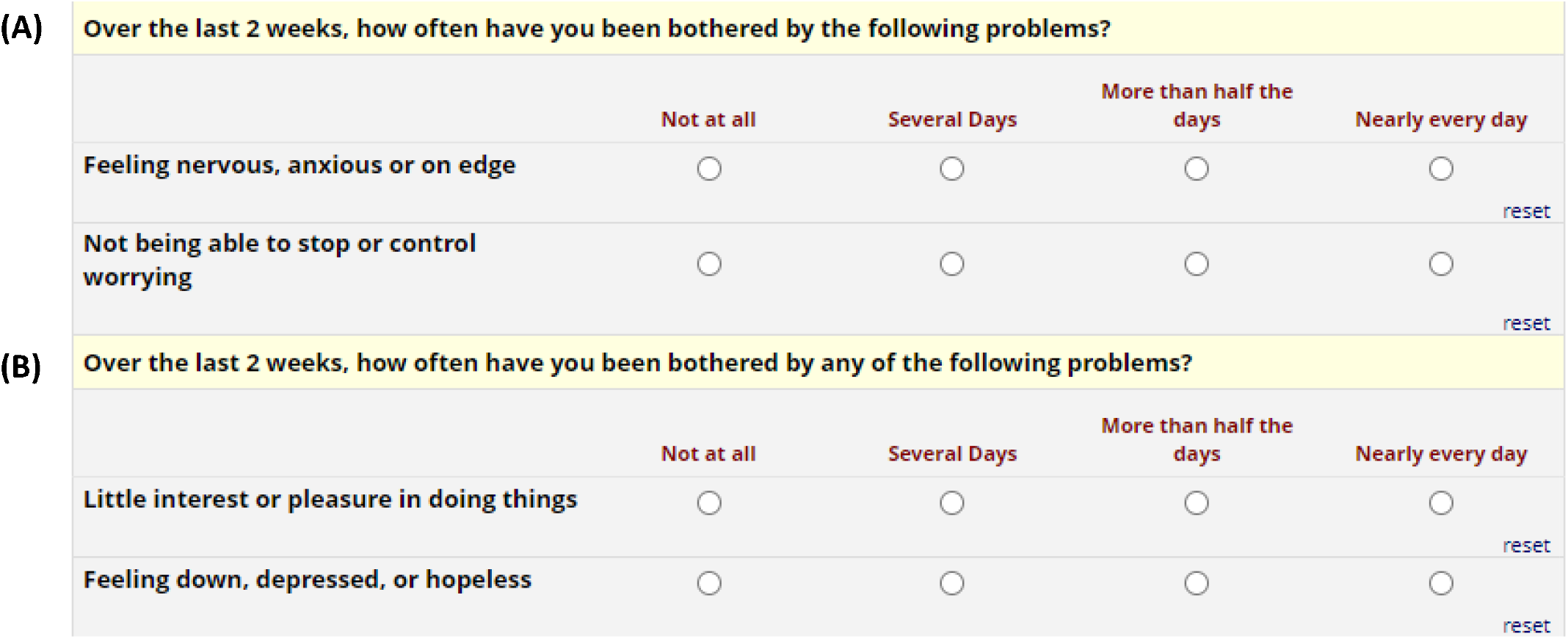
GAD2 (A) and PHQ2 (B)

## Notes

**Sources of Funding:** This work was supported by: NHLBI 1R01HL163024 (A.A.P), NHLBI K01HL140261 (A.A.P), CF Foundation PEZZUL20A1-KB (A.A.P), and Carver College of Medicine COVID-19 Grant (A.A.P), R01HL148758 (J.L.C).

### Competing Interest Statement

The authors have declared no competing interest.

### Funding Statement

This work was supported by: NHLBI 1R01HL163024 (A.A.P), NHLBI K01HL140261 (A.A.P), CF Foundation PEZZUL20A1-KB (A.A.P), and Carver College of Medicine COVID-19 Grant (A.A.P), R01HL148758 (J.L.C).

### Author Declarations

IRB of the University of Iowa gave ethical approval for this work.

